# Schizophrenia versus Healthy Controls Classification based on fMRI 4D Spatiotemporal Data

**DOI:** 10.1101/2025.01.26.25321159

**Authors:** Tieu Hoang Huan Pham, Yihren Wu, Toshikazu Ikuta, Majnu John

## Abstract

A wide array of machine learning approaches have been employed for differentiating patients with mental health disorders from healthy controls using neuroimaging data. However, almost all such methods have been applied on inputs based on connectivity matrices or features derived from the neuroimaging data. Only a few papers recently have considered such classification based on the original voxel-based spatiotemporal data. In this paper, we report the performance of a few cutting edge machine learning algorithms on voxel-based fMRI data to classify healthy controls and patients with schizophrenia. The methods that we employed included convolutional neural networks, convolutional recurrent neural networks with long short-term memory and a transfer learning approach for classification based on Wasserstein generative adversarial networks. In order to reduce the computational burden to fit in with available hardware, we had to reduce the original 4-dimensional data to 3-dimensional inputs for almost all architectures. Our results indicate that the relatively simpler architecture based on convolutional neural networks showed reasonable unambiguity in grouping patients from healthy controls. In contrast, the performance of the other two more complex architectures that we employed were comparatively poorer.

## 1. Introduction

Neuroimaging techniques such as magnetic resonance imaging (MRI) and positron emission tomography (PET), with their perceived potential as brain-based biomarkers, have been utilized in hundreds of clinical research studies over the past several decades. The interest and utilization of neuroimaging exponentially increased after the development of functional MRI (fMRI) and related statistical software in the 1990s [1,2]. The high dimensionality of fMRI, with data from hundreds of thousands of voxels at spatial resolution in the millimeter level (3 mm^3^), makes it a potential tool for exploring detailed contents of a person’s mental state [3,4]. Recent upgrades to MR technologies, improving the spatial precision to the submillimeter level, increase their ability even further to resolve minute details in brain structure and function [5]. Imaging biomarkers hold the promise of personalizing medicine by assisting in the understanding of the neural underpinnings of brain disorders as well as enhancing the diagnostic information for mental illnesses. In addition to the prospects of revealing the pathophysiology of diseases, neuroimaging biomarkers such as fMRI have been traditionally considered for detecting traits as well as prediction and classification of patients from healthy controls.

Machine learning has played a significant role in elucidation of structural and functional anatomy as well as prediction and classification of mental health patients versus healthy controls based on MRI data [6]. Kernel-based methods such as Support Vector Machines (SVM) were applied for classification based on neuroimages [7,8] soon after they were introduced to the machine learning research community [9] and later became widely used in the field because of their capability to learn nonlinear boundaries [10]. Ensemble machine learning approaches such as Random Forests [11] were also utilized from very early on [12] because of nice properties such as generalization capacity due to relatively lower number of parameters and robustness towards outliers. In the past decade or so, the popularity of deep learning led to several fruitful applications in the neuroimaging field as well [13–15]. Specialized subtypes of deep learning such as convolutional neural networks (CNNs) which are capable of hierarchical feature extraction from multidimensional data arrays such as natural images, and recurrent neural networks (RNNs) which are capable of processing sequential data have also been utilized for neuroimaging applications. Avbersek and Repovs [16] provides a survey of neuroimaging papers that have utilized deep learning, CNN and RNN architectures. Generative approaches such as generative adversarial networks (GANs) have also found application for neuroimaging modalities, primarily for data augmentation, generating images and for translating between imaging modalities [17–19].

Although a variety of currently available machine learning algorithms have been applied in several fMRI studies as mentioned above, almost all the approaches have applied the algorithms on connectivity matrices or features extracted from the original voxel-based data. The raw voxel-based fMRI spatiotemporal data is 4-dimensional (4D, that is four axes).

It consists of 3D images (2D slices stacked together) of blood oxygenation level dependent (BOLD) signals captured over time. Thus 3 spatial dimensions and time as the 4th dimension. Feature extraction or connectivity matrix calculation results in reduced dimensionality and information loss. For example, connectivity matrices rely on regions of interest (ROI) based approaches where the large number of voxels ( 300K) in the original data is grouped together, anatomically or statistically via clustering, into a few hundreds of brain regions and correlations calculated among these regions. Such data reduction may have advantages from computational and interpretation perspectives, but is not necessary when prediction is the goal and when using the current state of the art machine learning algorithms which can handle very high-dimensional data. In fact, keeping the input data in the original format may help the machine learning algorithms to learn on its own the inter-relationships among the voxel-based data and extract features relevant for prediction, and thus taking full advantage of the potential of such algorithms. It has also been observed that traditional convolutional neural network-based approaches that were originally developed for training based on natural images may not be ideal for connectivity matrices. Recently, graph convolutional network (GCN) and its adaptations and variations have been considered for fMRI connectivity neighborhoods [20]. Although better suited for connectivity matrices and promising, such methods have all the inherent limitations of the GCNs in general such as sensitivity to noise in the data and graph structure vulnerability.

Schizophrenia is a progressive neuropsychiatric disorder with enormous personal suffering, disability, family burden, premature death, and societal cost [21,22]. Standard diagnostic procedure for schizophrenia based on DSM-5 can be costly both in terms of time and resources and often lacks objectivity. Recently, several studies have explored the potential of objective tools for schizophrenia diagnosis, especially using machine learning algorithms. In this paper, we present results for schizophrenia-versus-healthy-controls classification based on raw 4D spatiotemporal fMRI data from a single site using a few neural network architectures.

There has been only a handful of papers that attempted detection of neural disorders using artificial neural networks to raw 4D fMRI data. Li et al [23] combined 3D CNNs with long short-term memory (LSTM) framework to detect Alzheimer’s disease using 4D fMRI data. Malkiel et al [24] also followed a 2-phase approach on various 4D fMRI datasets where in the first phase their algorithm obtained vectors from 3D brain images and then conducted reconstruction-based pre-training steps before propagating them through a multi-layer transformer network. Nguyen et al [25] presented a novel architecture for spatiotemporal fMRI 4D data that used residual convolutional neural networks for spatial feature extraction and self-attention mechanisms for temporal modeling. Kim et al [26] presented a memory and computation-efficient Swin Transformer architecture which implemented a 4D window self-attention mechanism and absolute positional embeddings. We present machine learning architectures based on convolutional neural networks, recurrent neural networks with convolutional LSTM layers and generative adversarial networks (GAN). Although, GANs are typically used as a generative algorithm, we adapt the GAN framework for classification.

The rest of the paper is organized as follows. In the next section we present an overview of details of underlying concepts related to the classifier architectures we employed in our analysis, namely, Wasserstein-GAN, CNN and RNN with convolutional LSTM. It is followed by a Results section where we present details of the data acquisition and pre-processing of fMRI images, as well as details about the training steps and model parameters involved in our classifier architectures. The results section also includes a subsection on performance comparison across our analytic strategies. We summarize and draw conclusions in the last section.

## 2. Methods

In this section, we present an overview of the methodological background concepts underlying the machine learning architectures used in our analyses. Although most of this is standard material, we present a review here mainly for the convenience of the non-machine-learning (that is, neuroimaging) specialist.

### 2.1 Classifier based on Generative Adversarial Network (GAN)

#### Overview of GAN concepts

GAN, an acronym for Generative Adversarial Networks, was a strategy proposed by Goodfellow et al [27], for generating a sample of pseudo-images with distributional properties very close to a sample of real images. In other words, any GAN’s goal is to generate ‘fake’ images that look very real. A GAN achieves its goal by making two component neural networks compete against each other. The two basic competing neural network architectures within a generic GAN architecture are labelled as ‘Generator’ and ‘Discriminator’. Wasserstein-GAN (W-GAN) is a variant of GAN that was proposed later in the literature to overcome certain limitations of the original GAN approach. Our overall strategy based on GAN (specifically, W-GAN) was to develop 1-class predictors for the schizophrenia group and for the control group separately, and then to combine the 1-class predictors to obtain a classifier for schizophrenia versus control categorization based on the 4D fMRI images. The 1-class predictors mentioned above are discriminator networks (critic-architecture in W-GAN terminology) that are components in two separate GANs - one GAN for generating images with the same underlying distribution as the images from the schizophrenia sample and the other GAN for generating images similar to images from the control sample. In order to better explain our overall approach, we describe briefly below the underlying concepts of GAN first and W-GAN next.

The type of inputs to the overall GAN architecture varies from those of its component architectures. Since the goal of the overall GAN architecture is to generate a set of pseudo-images similar to a given set of images, the input to the GAN will be just one sample of images either from the Schizophrenia group or from the control group. That is, we have separate GANs for the schizophrenia sample and the control sample. For clarity, we may notate the GAN that takes the Schizophrenia sample as input as GAN_SCZ_, and similarly assign the notation GAN_CTRL_ for the GAN that takes the control sample as input. Since the underlying concepts and the architectural details are the same for GAN_SCZ_ and GAN_CTRL_, for convenience at times we may drop the subscripts when referring to either of them. The Generator network takes data sampled from a random distribution, typically Gaussian distribution, as input and outputs fake-image data closely resembling the original input to the GAN. Thus, the outputs from the corresponding generators in GAN_SCZ_ and GAN_CTRL_ will respectively resemble the images from the schizophrenia and control samples. The Discriminator of any GAN is a 2-class classifier which takes in as input a pooled dataset consisting of fake images produced by the Generator and real images from the input sample for the overall GAN. Thus, at any given training or testing instance, the input for the Discriminator of the GAN will be a fake or real image and the output will be a label categorizing the input as either fake or real.

During training, the goals of the Generator and Discriminator are adversarial. Discriminator tries to identify fake images from real images even if the fake images look very much alike real images while as the Generator tries to create images that look real enough to trick the Discriminator. Because of these opposite goals, the mathematical objective functions for the two component networks in GAN are different while training, and thus GAN cannot be trained like a regular network. Each training iteration for GAN is divided into two phases. At the very beginning of the training – that is, at the 0^*th*^ iteration, the Generator is employed to produce a batch of fake images. At any given iteration, the Discriminator is trained in the first phase and the Generator in the second phase. The first phase starts with creating an input-batch for the Discriminator by sampling a set of real images from the overall-GAN’s input training sample (thus a set of images from the schizophrenia group in the case of GAN_SCZ_ and from the control group in the case of GAN_CTRL_) and adding an equal number of fake images produced by the Generator to complete the input-batch. A corresponding target vector is created by labeling the fake images as 0 and the real images as 1, and then the Discriminator is trained like any other classifier-network using binary cross-entropy as the loss function. The key point to note is that in the first phase, only the weights of the Discriminator-network are updated. In the second phase, prior to training the Generator, another batch of fake images are produced first. In this latter phase, the weights of the Discriminator are frozen (that is, training for the Discriminator suspended) and only the weights of the Generator are updated. We trick the Discriminator by feeding it only the pseudo-images produced by the Generator at the beginning of the second phase but labeling all of them falsely as 1, thus falsely indicating them as real images. This trick forces the Generator to produce pseudo-image samples with properties very close to the real samples such that they will appear real to the Discriminator. However, as training progresses, the Discriminator gets better at correctly identifying fake images which enforces the Generator to produce more convincing fake images at the second phase of each iteration. Looking at the GAN training from a deeper perspective, the Generator does not directly work with real images any time during training but slowly learns to produce convincing fake images via gradients flowing back through the Discriminator. In other words, information about the real images are contained in the secondhand gradients that are passed on to the Generator from the Discriminator and because of this gradient flow, the Generator’s capability as real-looking-image-creator increases significantly at later stages of the training by when the Discriminator’s capability as a classifier is substantially increased. For our analysis, the main point to keep in mind is that by the end of training the Discriminator in GAN_SCZ_, for example, will have become a sufficiently good classifier to discriminate between a real image from the schizophrenia sample and a pseudo-image which closely resembles a real image from the same sample, and hence if we use such a trained Discriminator on an input sample consisting of images from both schizophrenia and control groups, it should hopefully lead to even better classification. A similar argument can be made of the trained Discriminator from GAN_CTRL_ as well, and so finally if we combine the predictive capabilities of discriminator-networks from both GAN architectures, we will potentially have a very effective classifier for schizophrenia versus control based on 4D fMRI images.

#### Brief Introduction to W-GAN and the rationale for using it

Although conceptually GAN’s scheme looks great, practically it is very hard to train GAN. Convergence issues and mode collapse are two common problems encountered while training GAN. Lack of convergence could result in the GAN architecture producing suboptimal images with mediocre quality. Mode collapse is a term used in literature to describe the scenario where the GAN model repeatedly produces images with the same pattern and qualities thereby failing to cover the full range of possible variations in the output images. The problems with the original GAN are essentially tied to the cost functions used while training. In the original GAN architecture, the objective functions used are binary cross-entropy loss. Although separate loss functions are employed for both the Discriminator component and the overall GAN optimization, they are both the same type, namely, binary cross-entropy. As a side note, recall that the Generator will only be trained via the overall GAN model which incorporates the Discriminant as well, and hence a separate loss function is not necessary for the overall GAN. It is often the case in practice that GAN optimizes the Discriminator much faster than its Generator. Once optimality for the Discriminator is attained, it can be shown mathematically that minimizing the GAN objective function is equivalent to minimizing the Jenson-Shannon divergence (JSD) between the data distributions of the real images and those estimated by the model [28, 29]. The above-mentioned fact can be mathematically expressed as follows (closely following and summarizing the details from [28]).

We denote the Discriminator as *D* : χ→ [0, 1] as a function defined o n the ( fMRI image) sample space χ. Here we think of the Discriminator as a function that assigns to any given sample image a probability (of it being real). We denote the Generator as a function *G*_*θ*_, specifically a neural network parametrized by (that is, with a vector of weights) *θ* from a ‘noise space’ Ƶ to the sample space χ. Let ℙ_*r*_ and 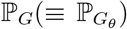 respectively denote the unknown distribution of the real images and distribution of pseudo-images outputted by the Generator, and we assume both distributions are continuous with densities *p*_*r*_ and *p*_*G*_. With this notation, the first phase of GAN training is to obtain a *D* that maximizes

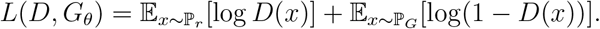

It is easy to note that [28] the optimal *D* is

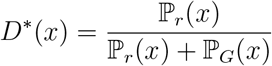

and that *L*(*D*^*^(*x*), *G*_*θ*_) = 2*JSD*(ℙ_*r*_|| ℙ_*G*_) −2 log 2 where *JSD*(ℙ_*r*_ ||ℙ_*G*_) denotes the Jensen-Shannon divergence which is a metric distance between ℙ_*r*_ and ℙ_*G*_. Hence in the second phase, minimizing *L*(*D, G*_*θ*_) as a function of *θ* is equivalent to minimizing *JSD*(ℙ_*r*_||ℙ_*G*_) when the Discriminator/Critic is optimal. Thus based on the theoretical considerations above one may think that a sound strategy would be to optimize *D* first so that we may use *JSD* as the cost function for optimizing *θ*. However, this does not work in practice primarily because of the vanishing gradients problem associated with *JSD* and secondarily because of the alternating nature of optimization between *D* and *θ*. The fact mentioned above related to vanishing gradients was theoretically shown in [28] under mild assumptions. Specifically, it was shown that

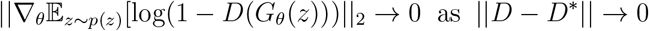

where *p*(*z*) denotes the density of the noise distribution from which the inputs for the Generator *G*_*θ*_ are sampled. In essence, the above theory confirms the fact that as the Discriminator gets better, the gradient of the Generator vanishes which makes GAN based on the original formulation extremely hard to train. To overcome this issue, a few alternatives were proposed including considerations of alternate cost functions. One proposed alternative was to use a different gradient step for the Generator,

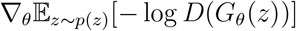

which effectively leads to optimizing a slightly different cost function. However, [2 8] theo-retically showed that although the above-mentioned alternative gradient approach does not suffer from the vanishing gradient problem, it could lead to massive variations in the gradients causing lack of convergence due to severely unstable updates during gradient descent. Yet another alternative suggestion in [28] was to add noise to the generated images, essentially to break the assumptions used in the previous theoretical statements. However even in this scenario theoretical issues occur, specifically tied to having *J SD* as the cost function when the Discriminator is optimal. Essentially, the issue arises from the mathematical fact that even with the tiniest amount of noise added to an original distribution, the *JSD* between the original and the noise-added distributions are maxed out although for all practical purposes the two distributions are very similar. All the above arguments led to considering an alternative distance metric, other than JSD, in the cost function. [28] proposed Wasserstein metric as the alternative and observed that it overcame all the limitations associated with JSD for noise-added data. Wasserstein’s distance, also known as the Earth Mover’s distance, between two distributions ℙ and ℚ on *X* is defined as

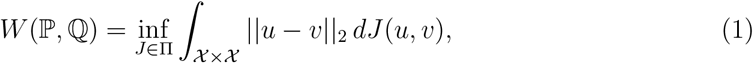

where ∏ denotes the set of all possible joint distributions on χ × χ with marginal distributions ℙ and ℚ. Quoting from [28], *W* (ℙ, ℚ) is “the minimum cost of transporting the whole probability mass of ℙ from its support to match the probability mass of ℚ on ℚ’s support.” Since the infimum based on (1) is highly intractable, an alternate formulation of the Wasserstein metric based on Kantorovich-Rubenstein (KR) duality [30] is more useful:

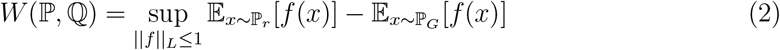

where the supremum is over all the 1-Lipschitz functions *f* : χ→ ℝ. With the KR formulation, the remaining hurdle is the selection of an appropriate 1-Lipschitz function that solves the optimization problem in (2). [29] proposed an approximation approach to obtain the optimal Discriminator by training a neural network *f*_*w*_ with weights *w*. This optimal Discriminator was called a ‘critic’ in the original paper since it was not trained to classify. The critic outputs a score function, rather than a probability, similar to the value function in Reinforcement Learning indicating how real the synthetic data is. Arjovsky proposed weight-clipping to enforce the Lipshchitz constraint. However, as noted in the original paper itself, weight-clipping can be problematic since if the clipping parameter is large, it takes a long term for weights to reach the limit while if the clipping is small, it leads to vanishing gradients when the number of layers is big or when batch normalization is not used. An alternative to weight-clipping proposed in a later paper by [31] was to use a gradient penalty as an application of the mathematical fact that a differentiable function is 1-Lipschitz if and only if it has gradients with norm at most 1 everywhere. Essentially, the idea is to constraint the gradient norm of the critic’s output with respect to its input by appending the penalty term

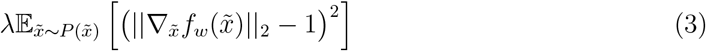

to the objective function in (2). Note that in (3), 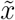 denotes a generic sample obtained from a straight line connecting a pair of samples from ℙ_*r*_ and ℙ_*G*_. Restriction to this interpolated sample is done for practical tractability reasons, and thus only a softer version of the constraint is implemented.

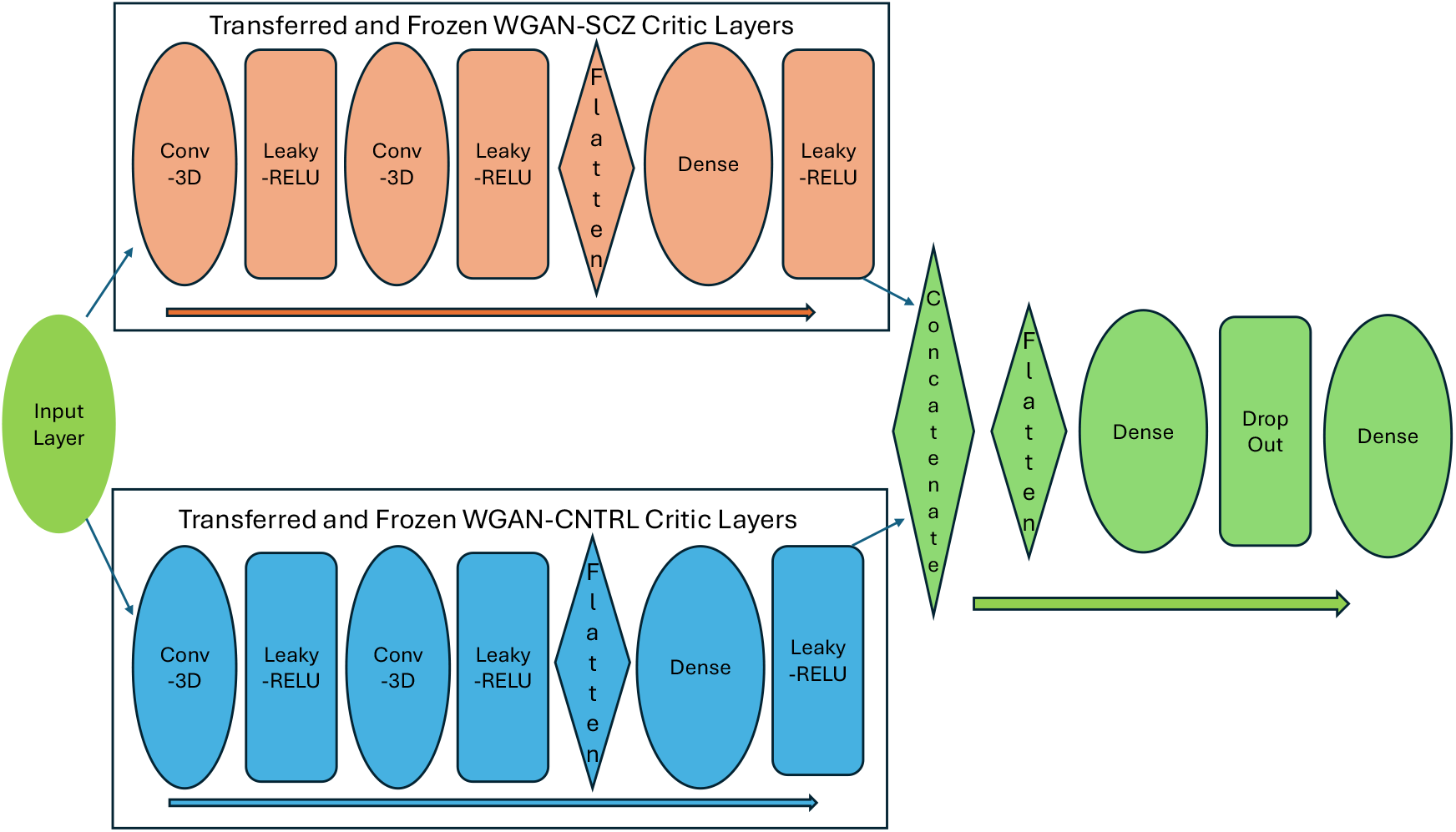

#### GAN based classification architecture via transfer learning

We use separate W-GAN’s for generative modeling, in parallel, of images from the patients with schizophrenia and from healthy controls. The critic network from within each W-GAN will be able to determine whether input data is from the corresponding assigned class or not. After the training for both the GANs are completed, the weights corresponding to the hidden layers within the critic network from the respective W-GANs are transferred into a main classifier model. Inside the main classifier model, the hidden layers from these separate GANs are arranged in parallel tracks. Note that the final layer from each critic model is not transferred - only the hidden layers are transferred. Once transferred, the weights for the critic’s hidden layers are frozen - that is, further training is not conducted on these transferred layers. Furthermore, the outputs from the two parallel pathways are concatenated and flattened before feeding into a fully connected layer and at the end passed through an output layer with sigmoid activation function for 2-class classification. Similar strategies have been previously employed in the literature in other contexts [32].

### 2.2 Classifier based on Convolutional Neural Network (CNN)

Over the past few decades convolutional neural networks (CNNs) have been widely used and highly successful in several applications including neuroimaging datasets. The key building block in a CNN is a convolutional layer which is a partially connected layer in contrast to fully connected layers seen in standard deep neural networks. CNNs with multiple convolutional layers have the ability to learn local as well as high-level patterns in data in a hierarchical manner by first extracting local, low-level features from raw input images and then increasingly more global and complex features in deeper layers. The input to a node in a convolutional layer is calculated via a convolution-operation conducted on the previous layer. The convolution-operation consists of first placing a window-grid (also known as a convolutional kernel or filter) locally over a section of the previous layer, secondly by conducting element-wise multiplication of value (weights) in the grid-cells of the filter and the corresponding values in the underlying cells in the previous layer over which the filter is placed, and finally by adding up all the multiplied values. Moving the filter-window horizontally and vertically, using the ‘stride’ parameter, over the previous layer yields the input values for different nodes in the convolutional layer. In order to expand the possibility of capturing wide variety of features, typically a large number of filters are used for each convolutional layer yielding input values for further nodes in the layer. Convolutional layers are often followed by a pooling layer with a max-pooling operation to reduce the dimensions and the computational load. Similar to the convolution-operation, the key step in the max-pooling operation is also done via placing a window-grid over the previous layer (this time, the previous layer is the convolutional layer). However, there are no weights associated with the cells in the window-grid for the max-pooling operation, and instead the maximum among the values in the previous layer framed by the window-grid, is extracted. A Batch-Normalization (BN) layer is often inserted in between convolutional layers to ameliorate the vanishing gradient problem during training.

### 2.3 Classifier based on Recurrent Neural Network (RNN)

Recurrent Neural Networks (RNNs) are neural networks specifically suited for learning patterns in sequential data such as time series. Simple RNNs are very limited with retaining short-term memory within the sequences. RNNs with Long-Short-Term-Memory (LSTM) cells are specialized models which are capable of learning longer term patterns in the data. At any given time point *t*, in addition to the data input **x**_*t*_, the two inputs that go into a LSTM cell are two state vectors **h**_*t*−1_ and **c**_*t*−1_ representing, respectively, the short-term state and long-term state from the previous time point. The outputs for the LSTM cell at time t are **ŷ**_*t*_, **h**_*t*_ and **c**_*t*_, where **ŷ**_*t*_ predicts the target vector **y**_*t*_, and **h**_*t*_ and **c**_*t*_ the updated state vectors. The equations governing the LSTM cell are

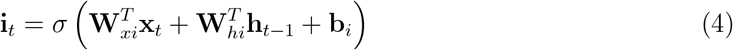

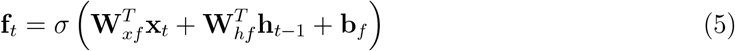

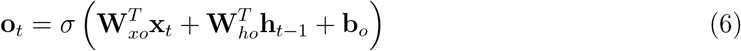

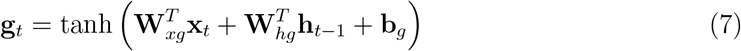

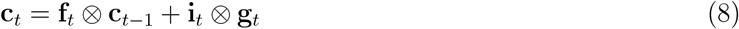

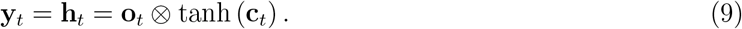

The first four equations above correspond to fully connected layers followed by an activation function - sigmoid in the case of the first three equations and hyperbolic tanget function in the case of the fourth equation. The **W**’s within the first four equations represent weight matrices for the corresponding layers and **b**’s represent the bias terms. The first three equations operate as gate controllers which depend on **x**_*t*_ and **h**_*t*−1_. As seen in the fifth equation above (that is, eq. (8)), **f**_*t*_ acts as a gate controller for the long term state **c**_*t*_, essentially as a decision-maker for which memories to be dropped from that state. Certain new memories are added to the long-term state **c**_*t*_ at time *t* based on the input **x**_*t*_ via its transformed version **g**_*t*_. The memory addition operation is controlled by the input gate **i**_*t*_. Finally, **o**_*t*_ represents the output gate which controls how the output **ŷ**_*t*_ and the short-term state **h**_*t*_ depends on the long-term state **c**_*t*_.

The standard form of LSTM mentioned above, which we call as FC-LSTM, is based on fully connected layers which do not take spatial correlation or any other spatial structure within the images into consideration. We used a version of LSTM known as the Conv-LSTM (short form for Convolutional LSTM) which has convolutional structures in both the input-to-state and state-to-state transitions (Shi et al), thereby accounting for the spatiotemporal relationships within the fMRI data. As with any fully connected networks, FC-LSTM flattens the inputs to 1-dimensional vectors leading to the loss of spatial information within the images. To avoid this drawback of FC-LSTM, the Conv-LSTM treats the inputs, long-term and short-term states, as well as the gates **i**_*t*_, **f**_*t*_ and **o**_*t*_, 3-dimensional tensors with the last two dimensions corresponding to the spatial dimensions (i.e. rows and columns of a 2-D array). In order to accommodate this change, the inputs, long-term and short-term states at time *t* are denoted as 𝔛_*t*_, 𝔠_*t*_ and 𝔥_*t*_, respectively. Each element in 𝔠_*t*_ and 𝔥_*t*_ is calculated based on values of the inputs and the past states of only its *local neighbors*, which is accomplished using a convolutional operator in the state-to-state and input-to-state transitions. Intuitively, with states conceptualized as hidden representations of moving objects, using larger kernels for convolution allows for capturing faster motion while as smaller kernels capture slower motions. As suggested in the original paper (Shi et al), zero padding is done before the convolutional operation to make sure that the states have the same number of rows and columns as inputs. With ‘*’ denoting the convolutional operator, the modified LSTM equations for ConvLSTM are

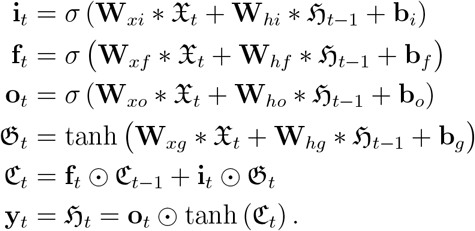

## 3. Results

In this section, details related to acquisition and pre-processing of fMRI images are presented first. Further subsections provide details about the training steps and model parameters involved in architectures used for schizophrenia versus healthy controls classification applied on the fMRI data set. A subsection on performance comparison across our analytic strategies is included at the end of the section.

### 3.1 Data Acquisition

The MRI images from the Center for Biomedical Research Excellence in Brain Function and Mental Illness [33] were obtained from Collaborative Informatics and Neuroimaging Suite (http://coins.mrn.org/). This data subset consisted of 182 individuals for whom resting state fMRI data were available. For the original study, subjects were excluded after screening if they had history of neurological disorder, history of mental retardation, history of severe head trauma with more than 5 minutes loss of consciousness, history of substance abuse or dependence within the last 12 months. The Structured Clinical Interview used for DSM Disorders (SCID) was used to determine the diagnostic information. Resting state echo planner image (EPI) volumes had 32 slices of 4 mm 64 × 64 matrix with 4-mm thickness (voxel size = 3 × 3 × 4 mm), with repetition time (TR) of 2000 milliseconds and echo time (TE) of 29 milliseconds. The maximum number of volumes per individual was 150 (5 minutes). High-resolution structural T1 volume was acquired as 176 sagittal slices of 256 mm × 256 mm with 1-mm thickness (voxel size = 1 × 1 × 1 mm, TR=2530 milliseconds and TE=3.25 milliseconds).

### 3.2 Data preprocessing steps

FMRIB Software Library (FSL) as well as Analysis of Functional NeuroImages were used for the standard preprocessing steps and statistical analyses of the data. he steps were similar to the ones described in [34]. As the first step in pre-processing of the images, skull-stripping, segmentation (gray matter, white matter and CSF) and registration to the MNI 2-mm standard brain was applied to the anatomical volume for each subject. The first 4 EPI volumes were removed. Transient signal spikes were removed by despiking interpolation. To correct head motion, the volumes were linearly registered to the then-first volume, through which 6 motion parameters and displacement distance between 2 consecutive volumes were estimated. Each of the resting state volumes was regressed by white matter and cerebrospinal fluid signal fluctuations as well as the 6 motion parameters. After smoothing with a 6-mm FWHM Gaussian kernel, the volumes were resampled, spatially transformed, and aligned to the MNI 2-mm standard brain space. Through this registration, 12 affine parameters were created between rs-fMRI volume and MNI152 2-mm space, so that a seed ROI can later be registered to each individual rs-fMRI space. To perform scrubbing where the volumes with excess motion are removed, as a displacement distance between 2 EPI volumes, the root mean square deviation was calculated from motion correction parameters, at an r=40-mm spherical surface using FSL’s rmsdiff tool [35, 36]. Volumes whose displacement distance exceeded the threshold (0.3 mm) were removed (scrubbed) from further statistical analyses [37].

We restricted our analysis to individuals for whom the fMRI time series had at least 90 time points, and after this reduction-step the total number of images available for the neural network based classification analyses was 141 with 59 images from patients diagnosed with schizophrenia and 82 images from age-matched healthy control subjects. For all the images that were retained in the final sample, the number of time points varied and hence zero-padding along the time axis was done to have consistent number of time points for all images. Further, images were normalized to have a range between −1 and 1 using the formula

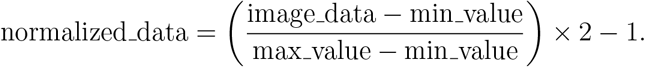

To make it computationally feasible for certain neural network architectures, one of the spatial dimensions of the input data was ‘collapsed’ so that the resulting input would have only two spatial dimensions. The details of these collapsing step are provided below where specific architectures are presented.

### 3.3 Overview of training strategies

Keras application programming interface [38] for Python with Tensorflow library [39] as the backend platform was used to built and train all the neural network models. Hardware played a not so insignificant role in our choices of certain model parameters. We used two hardware systems, which we label as Hw1 and Hw2 for further reference in the manuscript.

Hw1 was a workstation with the following specifications (CPU: Intel(R) Xeon(R) w5-2465X, 16 cores; RAM: 125 GiB (gibibytes); GPU: Nvidia RTX A6000 with 48 GB (gigabytes) of VRAM). Hw2 was a commercially available cloud computing resource (AWS Lambda) with on-demand gpu instances and clusters. We used a particular configuration called ‘gpu 8x a100 80gb sxm4’ which had 240 cores for the CPU and 1.9 TB for RAM available. Eight Nvidia A100 graphical processing units were pooled together to give 80GB of GPU memory (VRAM).

Random shuffle cross-validation was used for assessing the performance of each model. Fifteen cross-validation iterations were used all models except Conv-3D-LSTM. At each cross-validation iteration, random sampling from the whole data set was conducted to obtain a training data set consisting of 50 samples from the schizophrenia group and 50 samples from the healthy controls group. In the classifier neural network models, 20% of the training sample (10 images each from schizophrenia and control groups) was randomly set aside for validation (i.e. evaluation) while training. From the samples remaining after training data selection, a separate test sample of size 18 (9 from the schizophrenia group and 9 from the healthy controls) was randomly selected for performance evaluation of the models. Performance measures such as sensitivity, specificity and accuracy were calculated for each cross-validation iteration and the mean and standard deviation values of these measures across 15 cross-validation iterations are reported.

### 3.4 Model parameters and other specifics of neural network architectures

#### 3.4.1 W-GAN and transfer learning based classification

Within each of the cross-validation iteration, we trained two separate W-GAN models on 10000 epochs for each class (that is, one W-GAN for schizophrenia sample and another for the healthy-control sample). Further transfer learning was used to build a classifier with layers extracted from the W-GANs. The extracted layers were arranged in parallel tracks and output from these pathways were concatenated and flattened before inputting into a fully connected layer. The transfer-learning based classifier was trained for another 300 epochs specifically for classification task with all the transferred W-GAN layers kept frozen. Because of the lengthy time (several weeks) taken for training the W-GAN models we had to work with hardware available in our institution rather than commercially available servers (Hw2), and because of the hardware limitations of Hw1, we had to reduce the spatial dimensions of the input images from 3 to 2. We implemented this spatial dimension reduction by summing up the values along one spatial axis. Since we did this reduction separately for the x-, y- and z-axes, we had 3 separate WGAN-transfer-learning-based classifiers which we label as WGAN-x-collapsed, WGAN-y-collapsed, and WGAN-z-collapsed, respectively. The original dimensions of the input images after pre-processing steps including zero padding were: t-dimension = 146, x-dimension = 91, y-dimension = 109, and z-dimension = 91. Thus after collapsing the respective spatial axes, the input dimensions for the x-collapsed, y-collapsed and z-collapsed models were respectively (146, 109, 91), (146, 91, 91) and (146, 91, 109).

However, with the above-mentioned input sizes, the W-GAN models mentioned below were beyond the computational capacity of the Hw1 hardware. Thus, in order to accommodate the hardware limitations of Hw1, we further resized the input size for all WGAN models to be of dimensions (84, 84, 72). Only the input structure differed but otherwise the architectural details were the same for all the three WGAN-transfer-learning-based models and hence we describe the architectural details generically below.

For the WGAN architecture used in our analysis, the generator model started with a dense layer that maps noise input of dimension equal to 100 to an initial 3D volume of shape 7 × 7 × 9 ×128. This volume was progressively upsampled through a pair of transposed convolutional layers, with 64 and 32 filters of kernel size 3, respectively. Activation function used after each transposed convolutional layer was the Leaky version of Rectified Linear Unit (LeakyReLU) with negative slope set to 0.01. A final upsampling with another transposed convolutional layer with a *tanh* activation function was used to reach the target shape of generated synthetic 3D MRI volumes. The critic model in our WGAN was built using convolutional layers with 64 and 128 filters of size 3, respectively, as the first two layers. Downsampling was accomplished in these convolutional layers through strides. The output from the second convolutional layer was reshaped into a one dimensional array and passed through a dense layers with 128 nodes. LeakyRELU with negative slope equal to 0.01 was used as the activation function for the convolutional and dense layers. The final layer outputted a single score indicating the realness of the input. The final dense layer in the transfer-learning-based classifier had 256 nodes. The outputs from this dense layer passed through a Rectified Linear Unit (ReLU) activation function and then through a dropout layer with 0.3 dropout probability to prevent overfitting.

A batch size of 10 was used for all the WGAN models. For the generator and critic training in WGAN models, as well as for the transfer-learning based classifier model, a stochastic gradient descent optimization scheme known as ‘Adam’ optimization algorithm based on adaptive estimation of the first-order and second-order moments, was used. The learning rate, and the exponential decay rate for the first and second order moments were respectively set to 0.0001, 0.5 and 0.99. A number between 0 and 1 was uniformly sampled for each epoch and set as the mixing coefficient for interpolating the real and fake images to create the input sample for the gradient penalty. *λ* = 10 was used as the penalization parameter attached to the gradient penalty.

#### 3.4.2 CNN based classification

As with the inputs for the WGAN models, to accommodate computational resource constrictions related to hardware (Hw1), spatial reduction for the inputs by summing up values along a spatial axes. Thus we had three CNN models labelled CNN-x-collapsed, CNN-y-collapsed and CNN-z-collapsed based on the spatial axis for which the values were summed, and input sizes for these models were respectively (109, 91, 146), (91, 91, 146) and (91, 109, 146) with first two elements in each triple indicating the dimensions of the retained spatial axes and the last element indicating the temporal dimension of the fMRI data. Each CNN network consisted of three convolutional layers with Rectified Linear Unit (ReLU) activation function and progressively smaller kernel sizes: 10 × 10 × 10, 5 × 5 × 5, and 3 × 3 × 3. Each convolutional layer was followed by a max-pooling layer with a 2 × 2 × 2 pool size and batch normalization to stabilize learning. After the last convolutional layer, the model flattened the output into a single vector. The vectorized output was then passed through three fully connected (dense) layers, each with 5000 units and ReLU activation, interspersed with dropout layers to mitigate overfitting. The dropout probability in each dropout layer was set to 30%. The final output layer used a sigmoid activation for binary classification. The model was compiled with the Adam optimizer and binary cross-entropy loss to optimize for classification accuracy. 30 epochs were used for training each CNN model.

#### 3.4.5 RNN based classification

We implemented two versions of RNNs based on convolutional LSTMs (ConvLSTM). One model that kept all the four dimensions of the fMRI data (i.e. all three spatial dimensions and the time dimension) which we label as Conv-3D-LSTM, and the model that took in inputs with only 2 spatial dimensions similar to our earlier WGAN and CNN models, which we label as Conv-2D-LSTM.

*Conv-3D-LSTM* : This RNN model was constructed with an input shape corresponding to the time dimension and 3D spatial dimensions of the MRI volumes, with a single channel per volume. The model begins with an input layer of shape (146,91,109,91,1). A 3-dimensional Conv-LSTM layer with 2 filters and a 3 × 3 × 3 kernel was used to capture the spatiotemporal patterns, while maintaining the original spatial resolution through zero padding. The Conv-LSTM outputs were processed through a pooling layer with a global average pooling operation applied independently to each time step of the sequence, followed by a dense layer with 8 units and ReLU activation. The final output layer, a dense layer with a sigmoid activation, was used for binary classification. The model was compiled using the Adam optimizer and binary cross-entropy loss and trained using 100 epochs. This particular model was the only one implemented using the hardware Hw2.

*Conv-2D-LSTM* : This RNN model was designed to process 2D spatio-temporal MRI data, with an input shape of (time-steps, spatial-dimension-1, spatial-dimension-2, 1). As in our previous WGAN and CNN models, one spatial dimension was removed by summing up the values along that dimension. Thus we had x-collapsed, y-collapsed and z-collapsed versions of the RNN model with input shapes (146, 109, 91, 1), (146, 91, 91, 1) and (146, 91, 109, 1), respectively. The RNN architecture commenced with a 2-dimensional Conv-LSTM layer, which had 16 filters and a 3 × 3 kernel size to extract temporal and spatial features simultaneously. The output was a sequence of feature maps over time. The Conv-LSTM layer used zero padding to maintain the spatial resolution throughout the time steps. A 2-dimensional pooling layer was then applied to perform global pooling independently at each time step, reducing each feature map to a single value per filter. The pooled outputs were passed through a dense layer with 256 units and ReLU activation to further reduce the feature dimensionality. Finally, a dense output layer with a sigmoid activation was used for binary classification. The model was compiled using the Adam optimizer with custom parameters (learning rate = 0.001, and the exponential decay rate for the first and second order moments 0.9 and 0.99, respectively) to control momentum during training, and binary cross-entropy is used as the loss function to optimize classification performance. 75 epochs were used for training each model.

#### 3.4.4 Comparison of the classifier models

The comparison of all the architectures employed in our analyses, based on various performance measures are presented in the tables below. The performance measures that we considered were accuracy, sensitivity and specificity.

In terms of accuracy, the best performance was exhibited by the CNN architecture with ≥70% accuracy for all three types of input (that is, x-collapsed, y-collapsed and z-collapsed). This result is interesting because CNN is the least complex among all the architectures considered in our analyses. CNN applied on all three types of input performed better than the other architectures applied on any type of input. Among the other two architectures (ConvRNN and WGAN-TL) applied on inputs with reduced dimensions WGAN-TL did better overall in terms of accuracy. ConvRNN’s performance when applied on 3D inputs was quite poor with accuracy rates near 50%. ConvRNN’s accuracy rate (66.7%) was much better when applied on 4D input. However, no valid conclusions can be drawn from this result related to 4D output with only 1 CV iteration.

In terms of sensitivity and specificity presented in tables 2 and 3, CNN did consistently well and ConvRNN did consistently poor with all three types of 3D inputs. Interestingly, WGAN-TL’s sensitivity rates were much higher compared to its overall accuracy rates. The overall accuracy rates for WGAN-TL were brought down by the much lower specificity values. This discrepancy between sensitivity and specificity for WGAN-TL suggests that among the two underlying WGANs, the one for 1-class disrimination between real and fake fMRI images from patients with schizophrenia does better than the corresponding one for healthy controls. Another noteworthy point is that variation seen for sensitivity and specificity values with CNN architecture were much lower compared to the corresponding variation for the other two architectures. Thus CNN’s performance was superior not just in overall mean performance rates but also better precision of the estimated accuracy rates. Although ConvRNN did poorly for 3D inputs, the sensitivity when applied on 4D images was quite high at 89%, but again, no valid conclusions may be drawn with this result based on just 1-CV iteration.

**Table 1:** Average Accuracy (*±* SD) across 15 CV iterations.

| Dimension collapsed | WGAN-TL | CNN | ConvRNN |
| --- | --- | --- | --- |
| **x-collapsed** | 55.93% ± 09.3% | 77.04% ± 7.8% | 52.96% ± 9.4% |
| **y-collapsed** | 58.89% ± 11.5% | 70.00% ± 8.1% | 50.74% ± 13.4% |
| **z-collapsed** | 60.00% ± 11.7% | 73.33% ± 13.8% | 44.44% ± 14.8% |
| **None (i.e. 4D input)\*** | - | - | 66.7% |

\*Only 1 cross-validation iteration

**Table 2:** Average Sensitivity (*±* SD) across 15 CV iterations.

| Dimension collapsed | WGAN-TL | CNN | ConvRNN |
| --- | --- | --- | --- |
| **x-collapsed** | 67.41% ± 30.4% | 73.33% ± 14.4% | 57.04% ± 30.8% |
| **y-collapsed** | 78.52% ± 20.8% | 64.44% ± 14.1% | 50.37% ± 28.4% |
| **z-collapsed** | 65.19% ± 26.8% | 66.67% ± 23.0% | 37.78% ± 31.9% |
| **None (i.e. 4D input)\*** | - | - | 88.9% |

\*Only 1 cross-validation iteration

**Table 3:** Average Specificity accuracy (*±* SD) across 15 CV iterations.

| Dimension collapsed | WGAN-TL | CNN | ConvRNN |
| --- | --- | --- | --- |
| **x-collapsed** | 44.44% ± 27.5% | 80.74% ± 11.5% | 48.89% ± 26.2% |
| **y-collapsed** | 39.26% ± 23.3% | 75.56% ± 13.4% | 51.11% ± 24.8% |
| **z-collapsed** | 54.82% ± 30.1% | 80.00% ± 11.3% | 51.11% ± 35.6% |
| **None (i.e. 4D input)\* %** | - | - | 44.4% |
| *Only 1 cross-validation iteration |  |  |  |

## 4 Conclusions and Brief Discussion

The overall goal of our study was to assess the performance of a few of the currently cutting edge machine learning algorithms in classifying correctly schizophrenia patients and healthy controls based on voxel-based data from fMRI images. That is, our goal was to see whether any of the machine learning algorithms will classify well based on voxel-based data rather than data based on connectivity matrices or features extracted from the original data. The machine learning architectures that we employed in our study were based on convolutional neural networks, convolutional recurrent neural networks and a transfer learning approach for classification based on Wasserstein generative adversarial networks. Although our initial intention was to work with 4-dimensional spatio-temporal voxel-based data from fMRI images, because of hardware limitations we had to reduce the dimensions and work with 3-dimensional input data to lower the computational burden. We did apply one architecture (convolutional recurrent neural network) on the original unchanged 4D spatiotemporal data. However, only 1 cross-validation iteration was employed with 4D input while as 15 iterations were used for all methods applied on 3D inputs.

Some of the architectures that we employed were fairly complex and required lengthy time and substantial resources for training. For example, our WGAN-TL approach trained two separate Wasserstein generative adversarial networks for 10000 epochs each for the images from schizophrenia and control groups. The convolutional recurrent neural network architecture that we applied to the full 4D spatiotemporal data required commercially available cloud computing clusters with 240 cores for the central processing unit and 80 gigabytes of graphical processing units for virtual random access memory. However, our results indicated that the performance of the relatively simple architecture based on convolutional neural networks was substantially better compared to the other more complex architectures that we employed. The convolutional neural networks that we employed on 3D data exhibited accuracy, sensitivity and specificity values near 75% thereby showing that it is possible to differentiate patients with schizophrenia from healthy controls with reasonable correctness using voxel-based data alone. Although the complex architectures that we utilized did not classify the two groups unambiguously, further study is required to see whether the performance of these architectures will be improved with better hardware.

## Data Availability

All data produced in the present study are available upon reasonable request to the authors

## Codes availability

Codes used for this paper are available at https://github.com/HuanPham5/SCZvsControlsFMRI

